# Coblation versus Bipolar Diathermy Hemostasis in Pediatric Tonsillectomy Patients: Systematic Review and Meta-Analysis

**DOI:** 10.1101/2020.09.13.20193557

**Authors:** Mohammad Karam, Ahmad Abul, Abdulwahab Althuwaini, Talal Alenezi, Ali Aljadi, Abdulredha Almuhanna, Abdulrahman AlNaseem, Abdulmalik Alsaif, Athari Alwael

**Affiliations:** School of Medicine, University of Leeds, UK; School of Medicine, University of Manchester, UK; Department of ENT, Consultant Otorhinolaryngology Head and Neck Surgery, Al Jahra Hospital, Jahra, Kuwait

**Author notes:** contributed equally as first co-authors (Mohammad Karam, Ahmad Abul). **Corresponding Author:** Mohammad Karam, Address: 129 Candle House, 1 Wharf Approach, Leeds, United Kingdom.

**Keywords:** Tonsillectomy, Coblation, Bipolar Diathermy, Post-operative Pain, Hemorrhage

## Abstract

**Objective:** To compare the outcomes of coblation versus bipolar in pediatric patients undergoing tonsillectomy.

**Methods:** A systematic review and meta-analysis were performed as per the Preferred Reporting Items for Systematic Reviews and Meta-analyses (PRISMA) Guidelines and an electronic search of information was conducted to identify all Randomized Controlled Trials (RCTs) comparing the outcomes of coblation versus bipolar in pediatric patients undergoing tonsillectomy. Intraoperative bleeding, reactionary hemorrhage, delayed hemorrhage and post-operative pain were primary outcome measures. Secondary outcome measures included return to normal diet, effects on tonsillar bed, operation time and administration of analgesia. Fixed and random effects models were used for the analysis.

**Results:** Seven studies enrolling 1328 patients were identified. There was a significant difference between coblation and bipolar groups in terms of delayed hemorrhage (Odds Ratio [OR] = 0.25, P = 0.0007) and post-operative pain (standardized mean difference [MD] = -2.13, P = 0.0007). Intraoperative bleeding (MD = -43.26, P = 0.11) and reactionary hemorrhage did not show any significant difference. For secondary outcomes, coblation group had improved outcomes in terms of administration of analgesia, diet and tonsillar tissue recovery and thermal damage. No significant difference was reported in terms of operation time.

**Conclusions:** Coblation is a superior option when compared to bipolar technique for pediatric patients undergoing tonsillectomy as it improves post-operative pain and delayed hemorrhage and does not worsen intraoperative bleeding and reactionary hemorrhage.

**Highlights:**

- Coblation tonsillectomy was less painful than bipolar diathermy.
- Coblation tonsillectomy was associated with less delayed hemorrhage than bipolar diathermy.
- Tonsillar tissue recovery, thermal damage and return to normal diet were better in the coblation group.

## Introduction

Tonsillectomy is one of the oldest surgical operations in medicine and one of the most common operations performed by otolaryngologists^1,2^. Some potential indications for pediatric tonsillectomy include recurrent tonsillitis, sleep apnea as well as PFAPA Syndrome (Periodic Fever, Aphthous Stomatitis, Pharyngitis, Adenitis) and it is therefore one of the most common operations during childhood ^3^. Several techniques are used to perform tonsillectomies, including blunt dissection, guillotine, bipolar diathermy dissection, laser dissection, and the more recent coblation method ^4^. Despite the range of available techniques, postoperative pain, primary or reactionary hemorrhage, and postoperative infection associated with the hemorrhage continue to present as the main post-tonsillectomy complications ^5^. Therefore, studies continue to debate the optimal tonsillectomy technique.

Coblation (cold ablation) is a relatively new tonsillectomy technique that has earned increased popularity due to the decreased postoperative pain and reduced intraoperative bleeding that comes with its use ^6^. Coblation is operated at surface temperatures (40-70°C) much lower than those of more traditional techniques ^7^. Instead of relying on heat, coblation applies radiofrequency energy to a conductive natural salt solution, forming a plasma membrane comprised of highly ionised particles that hold enough energy to break the molecular bonds holding the tissue, thereby safely removing the target tissue ^7-9^. The use of coblation eliminates the risk of causing thermal damage that comes with heat and minimizes necrosis of surrounding healthy tissue, therefore resulting in minimal pain and faster recovery ^8,9^.

Bipolar diathermy is an electrosurgery technique that functions by passing an alternating current at a high-frequency through a pair of forceps to cut the tissue and coagulate the blood vessels^10^. Compared to monopolar diathermy, bipolar diathermy provides more control over the targeted area and uses less energy, thus causing less damage^10^. The use of bipolar diathermy to perform tonsillectomy is first described in a paper^11^ written in 1994. Bipolar diathermy tonsillectomy is proven to be safe for both adults and children^11,12^. Although the pain and morbidity rates are similar to other tonsillectomy techniques like cold dissection tonsillectomy, the bipolar diathermy technique provides significant advantages such as shorter operative time and lower blood loss levels^11, 12^.

Several published studies assessed the effectiveness of coblation compared with bipolar diathermy techniques in pediatric patients undergoing tonsillectomy^13-19^ including a previous meta-analysis comparing coblation to several techniques used in tonsillectomy^20^. However, this is the first study to perform a meta-analysis on these two specific techniques focusing on the pediatric population.

## Methods

A systematic review and meta-analysis were conducted as per the Preferred Reporting Items for Systematic Reviews and Meta-Analyses (PRISMA) guidelines^21^.

### Eligibility criteria

All randomized control trials and observational studies comparing coblation versus bipolar diathermy hemostasis techniques for tonsillectomy were included. Coblation was the intervention group of interest and bipolar diathermy was the comparator. The study also included patients having bipolar scissors, bipolar forceps as well as cold steel and bipolar diathermy hemostasis as the comparator. All patients were included irrespective of gender or co-morbidity status as long as they belonged to either a study or control group and were pediatric patients. All case reports, cohort studies without a comparison group, and studies not written in English were excluded. The study also excluded the following techniques: bipolar molecular resonance coagulation, tonsillotomy, unipolar or monopolar diathermy, adenotonsillectomy and conventional or traditional tonsillectomy without explicitly stating bipolar diathermy as the primary method for hemostasis.

### Primary Outcomes

The primary outcomes are intraoperative bleeding, reactionary hemorrhage (within 24 hours after the operation), delayed hemorrhage (bleeding after 24 hours of the operation) and post-operative pain on day 7.

### Secondary Outcomes

The secondary outcomes included return to normal diet, effects on tonsillar bed (degree of healing in tonsillar fossae and thermal damage to tonsillar tissue), operation time and administration of analgesia.

### Literature search strategy

Two authors AA and MK independently searched the following electronic databases: MEDLINE, EMBASE, EMCARE, CINAHL, and the Cochrane Central Register of Controlled Trials (CENTRAL). The last search was run on the 12^th^ of April 2020. Thesaurus headings, search operators and limits in each of the above databases, were adapted accordingly. In addition, World Health Organization International Clinical Trials Registry (http://apps.who.int/trialsearch/), ClinicalTrials.gov (http://clinical-trials.gov/), and ISRCTN Register (http://www.isrctn.com/) were searched for details of ongoing and unpublished studies. No language restrictions were applied in our search strategies. The search terminologies included “coblation”, “bipolar”, and “tonsillectomy”. The bibliographic lists of relevant articles were also reviewed.

### Selection of Studies

The title and abstract of articles identified from the literature searches were assessed independently by two authors (AA and MK). The full texts of relevant reports were retrieved and articles that met the eligibility criteria of our review were selected. Any discrepancies in study selection were resolved by discussion between the authors.

### Data Extraction and Management

An electronic data extraction spreadsheet was created in line with Cochrane’s data collection form for intervention reviews. The spreadsheet was pilot tested in randomly selected articles and adjusted accordingly. Our data extraction spreadsheet included study-related data (first author, year of publication, country of origin of the corresponding author, journal in which the study was published, study design, study size, clinical condition of the study participants, type of intervention, and comparison), baseline demographics of the included populations (age and gender), and primary and secondary outcome data. The authors cooperatively collected and recorded the results and any disagreements were solved via discussion.

### Data synthesis

Data synthesis was conducted using the Review Manager 5.3 software. The extracted data was entered into Review Manager by three independent authors. The analysis involved used was based on fixed and random effects modelling. The results were reported in forest plots with 95% Confidence Intervals (CIs).

For dichotomous outcomes, the Odds Ratio (OR) was calculated between the 2 groups. The OR is the odds of an event in the coblation group compared with the bipolar group. An OR of greater than 1 for the delayed hemorrhage would favor the coblation group, an OR of less than 1 would favor the bipolar group, and a OR of 1 would favor neither groups.

For continuous outcomes, the Mean Difference (MD) was calculated between the 2 groups. A positive MD for the post-operative pain score by day 7 and intraoperative bleeding would favor the coblation group, a negative MD would favor the bipolar group, and an MD of 0 would favor neither groups.

### Assessment of Heterogeneity

Heterogeneity among the studies was assessed using the Cochran Q test (χ^2^). Inconsistency was quantified by calculating I^2^ and interpreted using the following guide: 0% to 25% may represent low heterogeneity, 25% to 75% may represent moderate heterogeneity, and 75% to 100% may represent high heterogeneity.

### Methodological quality and risk of bias assessment

Multiple authors independently assessed the methodological quality as well as the risk of bias for articles matching the inclusion criteria. For randomized trials, the Cochrane’s tool for evaluating the risk of bias was used. Domains assessed included selection bias, performance bias, detection bias, attrition bias, reporting bias, and other possible sources of bias. Randomized controlled trial (RCT) studies are classified into low, unclear, or high risk of bias. For non-randomized studies, the Newcastle-Ottawa Scale was used^22^. It uses a star grading system to assess studies in terms of three domains: selection, comparability, and exposure. The total maximum score for each study is nine stars. The overall rating of either good, fair or poor quality was based on the Agency for Healthcare Research and Quality (AHRQ) standards^22^.

## Results

### Literature search results

The search strategy retrieved 546 studies and after a thorough screening of the retrieved articles using the inclusion and exclusion criteria, the authors identified seven studies in total which met the eligibility criteria (Figure 1).

**Figure 1:**
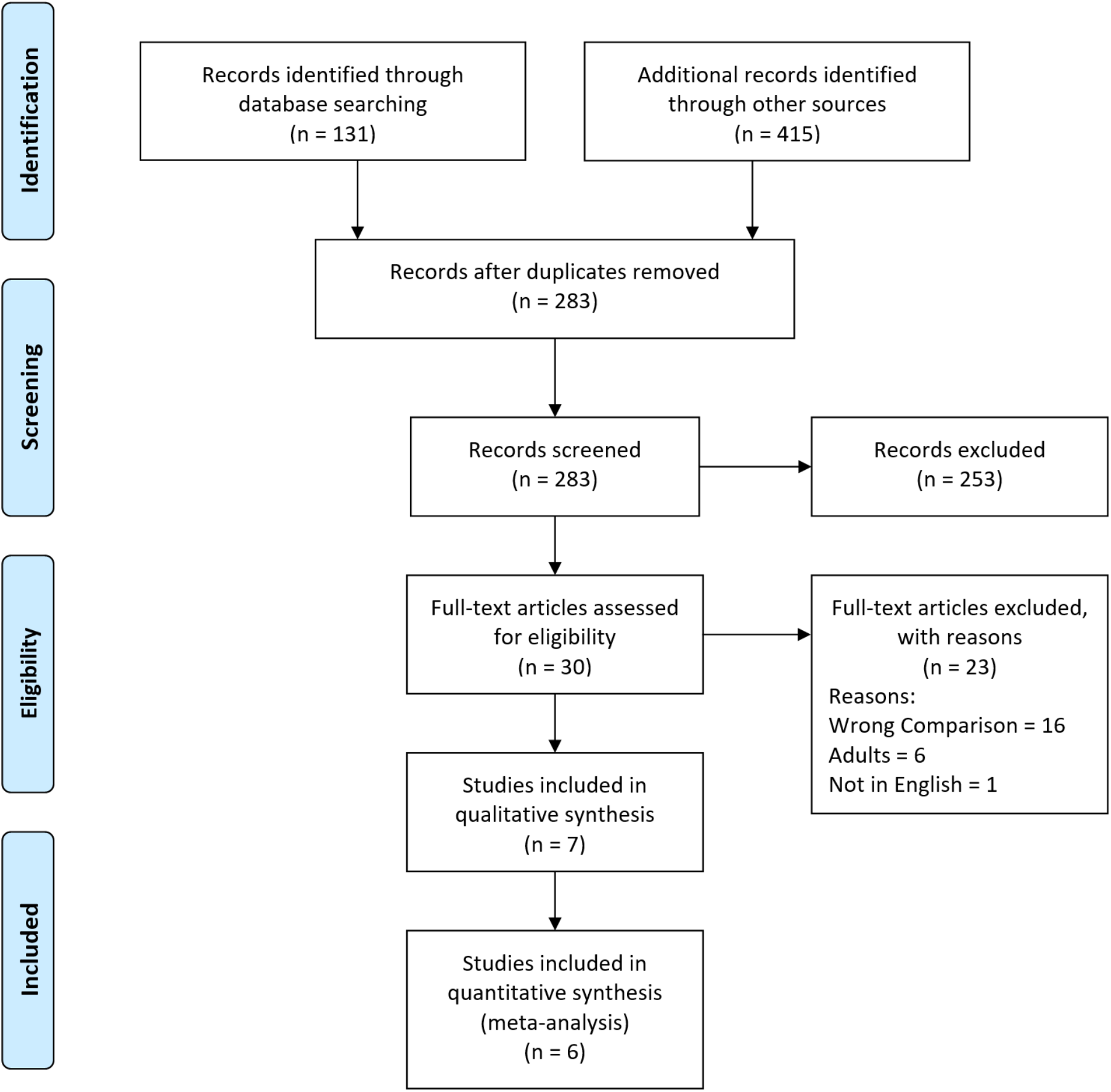
Prisma Flow Diagram. The PRISMA diagram details the search and selection processes applied during the overview. PRISMA: Preferred Reporting Items for Systematic Reviews and Meta-Analyses.

### Description of Studies

Table 1 summarises the baseline characteristics of included studies published from 2001 to 2018. All included population were pediatric patients under the age of 18 who underwent tonsillectomy.

**Table 1.** Baseline Characteristics of the Included Studies. NR: not reported.

| Study (Year) | Country, Journal | Study Design | Sex (Male:Female) | Mean Age ± SD (Range) | Total Study Sample (Control:Intervention) | Interventions Compared |
| --- | --- | --- | --- | --- | --- | --- |
| Temple et al. (2001) | International Journal of Paediatric otorhinolaryngology, UK | RCT‡ | 19:19 | 5.6 years | 38 (18:20) | Standard bipolar versus coblation tonsillectomy |
| Mitic et al. (2007) | Clinical Otolaryngology, Norway | RCT | NR† | NR (4-12 years) | 40 (20:20) | Coblator or steel dissecting instruments versus bipolar diathermy |
| Parker et al. (2009) | Clinical Otolaryngology, UK | RCT | 35:44 | 8.2 years (4-15 years) | 70 (35:35) | Cold steel dissection with bipolar hemeostasis versus coblator dissection |
| Roje et al. (2009) | Collegium antropologicum, Croatia | RCT | 41:31 | 6 years (3-16 years) | 72 (36:36) | Conventional cold steel tonsillectomy with bipolar diathermy coagulation versus coblator |
| Konsulov et al. | International Journal of Otorhinolaryngology, Bulgaria | Prospective Comparative Study | NR | NR (3-18 years)<br>Coblation: $8.16 \pm 4.74$<br>Bipolar: $6.87 \pm 3.01$ | 60 (30:30) | Coblator II system ArthroCare (Smith and Nephew) Evac 70 wand for extracapsular dissection vs. extracapsular blunt dissection with bipolar diathermy |
| Belloso et al. (2010) | The Laryngoscope, UK | Cohort | NR | Coblation: $7.76 \pm 3.56$ years<br>Bipolar: $7.60 \pm 3.44$ years | 1008 (526:482) | Coblation or blunt dissection versus bipolar diathermy hemostasis |
| Bhardwaj et al. (2018) | Indian Journal of Otolaryngology and Head & Neck Surgery, India | RCT | NR | NR (4-14 years) | 100 (50:50) | Coblation Assissted versus Bipolar Diathermy |

† NR: Not Reported
‡ RCT: Randomized Control Trial

#### Temple et al. 2001

Temple et al. conducted a single center prospective RCT that included 38 pediatric patients listed for a routine tonsillectomy with a history of chronic tonsillitis or obstructive tonsils. Patients were randomized via closed opaque envelope technique to either have bilateral coblation tonsillectomy (using an ArthroCare CoVac™ 70 ArthroWand®) or bilateral standard bipolar dissection tonsillectomy. All tonsillectomies were extracapsular.

#### Mitic et al. 2007

Mitic et al. performed a single center prospective RCT that included 40 patients with standard indication for tonsillectomy. Randomizing was achieved by closed opaque envelope technique to either have coblation tonsillectomy or dissection tonsillectomy with bipolar diathermy hemostasis. Bipolar dissection was done using standard technique (set at 4/10 and 50 Watts power). All tonsillectomies were extracapsular.

#### Parker et al. 2009

Parker et al. performed a single center prospective RCT that included 60 pediatric patients undergoing tonsillectomy by either cold steel dissection or coblator dissection. The trial was double blinded and a computer-generated random sequence, in sealed opaque envelopes, was used for allocation of procedure technique. All tonsillectomies were extracapsular.

#### Roje et al. 2009

Roje et al. performed a single center, prospective RCT that included 72 pediatric patients listed for tonsillectomy. Randomization was fulfilled by using computer-generated random numbers that separated the patients into one of two groups that would undergo either coblation tonsillectomy or conventional cold-steel tonsillectomy with bipolar diathermy. All tonsillectomies were extracapsular.

#### Belloso et al. 2010

Belloso et al. performed a single center prospective observational cohort study which included 1008 participants from July 2001 to January 2003; 526 patients received a coblation tonsillectomy and 482 patients received a blunt dissection tonsillectomy. Participants data was extracted from a tonsillectomy audit. All tonsillectomies were extracapsular. Bipolar dissection was done using standard technique (set at 4/10- and 50-Watts power).

#### Konsolov et al. 2017

Konsolov et al. carried out a single center prospective cohort study which included 60 children aged 3-18 years. The children were divided into two equal groups. One group received traditional blunt dissection with bipolar diathermy hemostasis and the other underwent coblation tonsillectomy using the ArthroCare CoVac™ 70 ArthroWand®. All tonsillectomies were extracapsular.

#### Bhardwaj et al. 2018

Bhardwaj et al. conducted a single center prospective RCT that included 100 pediatric patients undergoing tonsillectomy. Patients were given random numbers and they got randomly allocated to one of two groups: one group undergoing bipolar diathermy and the other undergoing coblation. Bipolar dissection was done using 12 watts of power. All tonsillectomies were extracapsular.

### Primary Outcomes

#### Bleeding

Three outcomes of bleeding were reported in the included studies, namely intraoperative hemorrhage, reactionary hemorrhage and delayed hemorrhage.

Intraoperative bleeding was reported in two studies included 132 patients as shown in Figure 2. The mean difference analyses showed no statistically significant difference however a trend is demonstrated favoring the coblation group (MD = -43.26, CI = -96.33 to 9.80, P = 0.11). A high level of heterogeneity was found amongst the studies (I^2^ = 99%, P <0.00001). Mitic et al. also reported less intraoperative bleeding in the coblation group (28.25 mL) than the bipolar group (62.25 mL). However, Hasan et al. reported no significant difference between both groups.

**Figure 2:**
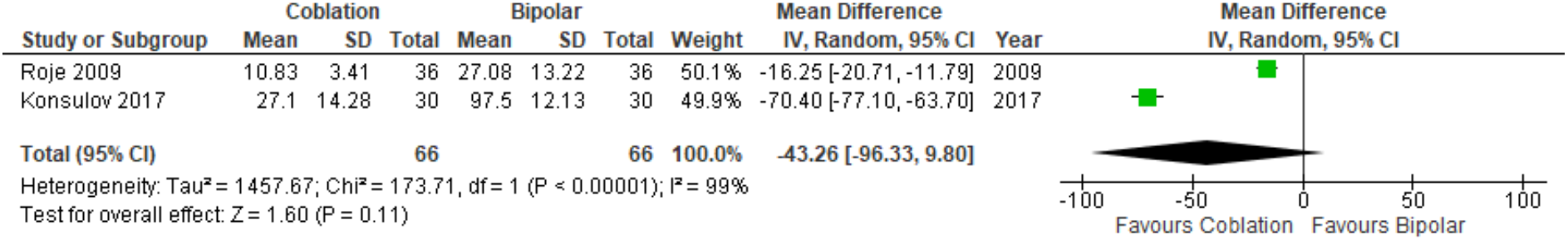
Forest Plot of Coblation versus Bipolar Tonsillectomy - Intraoperative Bleeding. Quantitative analysis showing the mean difference in the intraoperative bleeding reported as median by Roje et al. (2009) and Konsulov et al. (2017).

Reactionary or primary hemorrhage was reported by Roje et al. who did not record any case of primary hemorrhage in both groups. However, Kunsulov et al. reported two cases of reactionary hemorrhage in the bipolar group, compared to none in those that underwent coblation tonsillectomy.

Delayed or secondary hemorrhage was reported in four studies enrolling 1210 patients as demonstrated in Figure 3. There was a statistically significant difference seen in the odd ratio analyses showing a lower rate of delayed hemorrhage for the coblation group (OR = 0.25, CI = 0.11 to 0.55, P = 0.0007). A low level of heterogeneity was found amongst the studies (I^2^ = 7%, P = 0.34).

**Figure 3:**
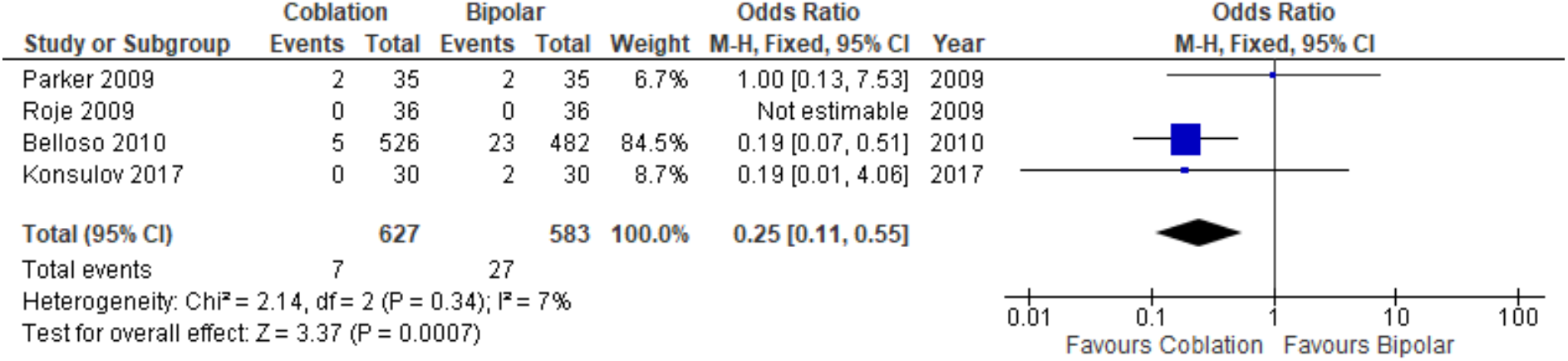
Forest Plot of Coblation versus Bipolar –. Delayed Haemorrhage. Quantitative analysis showing the odd ratio in delayed haemorrhage reported by Parker et al. (2009), Roje et al. (2009), Belloso et al. (2010) and Konsulov et al. (2017).

#### Post-operative Pain by Day 7

In Figure 4, post-operative pain by day 7 was reported using different pain scales in three studies enrolling 200 patients. There was a statistically significant difference seen in the standardized mean difference analyses showing less pain for the coblation group (standardized MD = -2.13, CI = -3.37 to -0.90, P = 0.0007). A high level of heterogeneity was found amongst the studies (I^2^ = 91%, P < 0.0001). Temple et al. also reported mean post-operative pain scores however did not include standard deviation; hence it wasn’t possible to quantitatively assess them in the forest plots. Temple reported a significant difference favoring coblation (p <0.0001).

**Figure 4:**
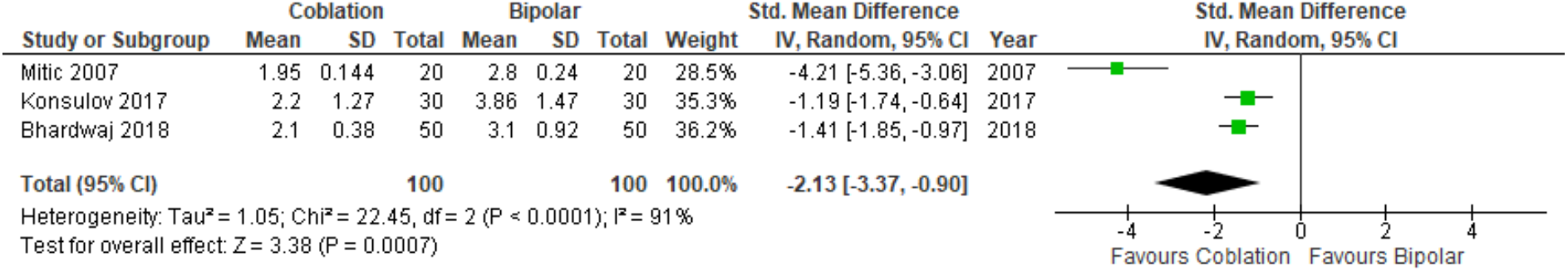
Forest Plot of Coblation versus Bipolar -. Post-operative Pain by Day 7. Quantitative analysis showing the odd ratio in delayed haemorrhage reported by Mitic et al. (2007), Konsulov et al. (2017) and Bhardwaj et al. (2018).

### Secondary outcomes

#### Return to Normal Diet

Three studies with two different types of assessment of diet were included. Temple et al. reported a statistically significant difference in the days it took to return to normal diet between the two groups, with an average of 2.4 days for patients who had coblation tonsillectomy versus an average of 7.6 days for patients who had routine bipolar dissection. On the other hand, Parker et al. reported no significant difference in the number of days taken by the two groups to return to normal diet, with a steady increase from day 6 onwards in both groups. Mitic et al. reported diet using a nutrition score during a 10-day postoperative period and found that there was a statistically significant difference between the score of patients who had coblation tonsillectomy and those who had dissection tonsillectomy across the 10 days, favoring the former.

#### Effect on Tonsillar Bed

A significant difference in the degree of healing in tonsillar fossae between the two interventions was reported by Temple et al., with nearly all coblation fossae healed 9 days post-operatively, while bipolar dissection patients had considerable slough. Roje et al. reported a statistically significant mean difference of the depth of thermal damage (t = - 40,1; p<0.001) to tonsillar tissue, where the coblation technique caused damage two times shallower than that caused by bipolar diathermy hemostasis (428.58 ± 47.4 um and 841.17 ± 39.7 um, respectively).

#### Operation time (min)

Operation time was defined as point of knife to skin contact according to Mitic et al.. This study reported a statistically insignificant difference between coblation and bipolar groups (26.6 min and 25.6 min, respectively).

#### Administration of Analgesia

Four studies have assessed analgesics administration in different ways. Parker et al identified less analgesic requirements by patients that underwent coblation tonsillectomy in the first 12 hours post-operatively. In addition, Roje et al and Konsulov agreed that patients undergoing coblation required lower number of days on analgesics for the coblation group compared to those that received the bipolar technique. Roje also identified that the coblation group required a lower number of analgesic applications (4 vs. 8). Mitic et al has reported a lower medication intake score for the coblation group however did not specify how the scale works. Other than Roje, the studies have mentioned using the same analgesics (ibuprofen and paracetamol).

### Methodological Quality and Risk of Bias Assessment

The Cochrane Collaboration’s Tool was used to assess the quality of the RCTs included in the study (Table 2). The Newcastle-Ottawa scale was used to assess the quality of the non-randomized studies (Table 3) which offers a star system for analysis^22^. The quality of the included non-randomized study was deemed to be high in selection and exposure but low in comparability. Overall, both studies were of good quality based on the AHRQ standards^22^.

**Table 2.** Bias analysis of the Randomized Trials using the Cochrane Collaboration’s Tool

| First Author | Bias | Authors' Judgement | Support for Judgement |
| --- | --- | --- | --- |
| **Temple et al. (2001)** | Random sequence generation (selection bias) | Unclear Risk | No information given |
|  | Allocation concealment (selection bias) | Low Risk | Closed opaque envelope technique |
|  | Blinding of participants and personnel (performance bias) | High Risk | Single surgeon – not blinded. |
|  | Blinding of outcome assessment (detection bias) | Low Risk | Outcome measurement is not likely to be influenced by lack of blinding |
|  | Incomplete outcome data (attrition bias) | High Risk | Standard deviation for pain is not reported |
|  | Selective reporting (reporting bias) | Unclear Risk | No information given |
|  | Other bias | Low Risk | Similar baseline characteristics in both groups. |
| **Mitic et al. (2007)** | Random sequence generation (selection bias) | Low Risk | A randomised sequence was generated by a statistician of the two interventions. |
|  | Allocation concealment (selection bias) | Low Risk | Allocation of treatment was envelope sealed by a statistician and was then opened in the surgery room in sequential order by a nurse. |
|  | Blinding of participants and personnel (performance bias) | High Risk | Surgeons were not blinded |
|  | Blinding of outcome assessment (detection bias) | Low Risk | Outcome assessment was done by parents and nurses whom all were blinded for the operation method. The data from parents and nurses were compared and no difference was identified. |
|  | Incomplete outcome data (attrition bias) | Low Risk | All questionnaires were complete. |
|  | Selective reporting (reporting bias) | High Risk | Return to normal diet mentioned in abstract was not reported. |
|  | Other bias | Low Risk | Similar baseline characteristics in both groups. |
| **Parker et al. (2009)** | Random sequence generation (selection bias) | Low Risk | Random sequence generated by envelope |
|  | Allocation concealment (selection bias) | Low Risk | Sealed opaque envelope technique |
|  | Blinding of participants and personnel (performance bias) | High risk | Single surgeon. |
|  | Blinding of outcome assessment (detection bias) | Low Risk | Double blinded study. The children, the parents and the nursing staff doing the pain assessments and prescribing the analgesia were not informed which technique had been used. The surgeon took no part in the pain assessments. |
|  | Incomplete outcome data (attrition bias) | Low Risk | Missing outcome data balanced in numbers across intervention groups, with similar reasons for missing data across groups |
|  | Selective reporting (reporting bias) | Low Risk | All outcome data reported |
|  | Other bias | Low Risk | Similar baseline characteristics in both groups. |
| **Roje et al. (2009)** | Random sequence generation (selection bias) | Low Risk | Computer-generated randomisation was used for selection of children and separating them into groups from a large ENT database. |
|  | Allocation concealment (selection bias) | Unclear Risk | No information given |
|  | Blinding of participants and personnel (performance bias) | High Risk | Single blinded (parents). Single surgeon not blinded |
|  | Blinding of outcome assessment (detection bias) | Unclear Risk | Parents assessing secondary outcomes did not know which procedure was used. |
|  | Incomplete outcome data (attrition bias) | High Risk | Participants lost to follow up (15 total- 7 coblation and 8 bipolar group) |
|  | Selective reporting (reporting bias) | Unclear Risk | Insufficient information. |
|  | Other bias | Low Risk | Similar baseline characteristics in both groups. |
| **Bhardwaj et al. (2018)** | Random sequence generation (selection bias) | Unclear Risk | Did not explain the method of obtaining the random numbers |
|  | Allocation concealment (selection bias) | Unclear Risk | Random numbers but not explicitly concealed |
|  | Blinding of participants and personnel (performance bias) | Unclear Risk | Insufficient information |
|  | Blinding of outcome assessment (detection bias) | Unclear Risk | Insufficient information |
|  | Incomplete outcome data (attrition bias) | Low Risk | No missing information |
|  | Selective reporting (reporting bias) | Low Risk | No published protocol but all the expected and pre-specified outcomes are reported |
|  | Other bias | Low Risk | No significant demographic difference between the 2 groups. |

**Table 3.** Newcastle-Ottawa Scale to Assess the Quality of Non-randomised Studies.

| Study | Selection | Comparability | Exposure |
| --- | --- | --- | --- |
| Belloso et al. (2010) | **** | * | *** |
| Konsulov et al. (2017) | **** | * | ** |

## Discussion

Coblation showed a superior effect when compared with bipolar diathermy in pediatric patients undergoing tonsillectomy in terms of delayed hemorrhage and postoperative pain, as shown by the results of the analyses. Although there were no significant differences between the coblation and bipolar groups in terms of intraoperative bleeding (P = 0.11) and reactionary hemorrhage, delayed hemorrhage was significantly (P = 0.0007) lower in the former group. Similarly, a significant (P = 0.0007) improvement in post-operative pain was noted for the coblation group. Regarding the between-study heterogeneity, it was low for delayed hemorrhage (I^2^ = 7%) and high for post-operative pain (I^2^ = 91%) and intraoperative bleeding (I^2^ = 99%), based on the assessment mentioned in the methodology.

Along with the outcomes mentioned above, the findings of this study reported several secondary outcomes that proved coblation to be a more effective technique than bipolar. Both tonsillar tissue recovery and thermal damage were significantly better in the coblation group. This correlates with analgesic administration whereby coblation required less doses compared to bipolar dissection. Generally, return to normal diet was quicker for the coblation group. There was no significant difference between the two groups in terms of operation time.

Currently, there is a debate in the literature with regards to the most efficient technique used for tonsillectomy in children^23^. The results of the current study are comparable to some studies where coblation tonsillectomy was found to be less painful than bipolar tonsillectomy in the immediate and overall postoperative period, which resulted in a swifter recovery and reduced analgesic requirements^24, 25^. A randomized control trial which included 80 adolescents found that pain and otalgia post-operatively was very slightly lower in the coblation group; this difference was, however, considered clinically insignificant^25^.

A more recent study in Iraq^26^, which compared intraoperative efficiency and postoperative recovery between bipolar electrocautery and coblation tonsillectomy in children, recorded statistically and clinically significant higher amounts of intra-operative blood loss in bipolar technique than using coblation (Bipolar 1.43 ml vs. Coblation 15.37 ml, P <0.001). Although this outcome conflicts with the results of this study, the coblation technique was associated with lower mean pain scores, which is the direction of this study^26^.

A study in the United States included 7,562 patients under 12 years of age compared the costs of treatment and management of children undergoing tonsillectomy by either coblation or electrocautery technique^27^. The study reported statistically significant lower costs for coblation compared to electrocautery surgery ($1,009 versus $1,162; P <0.0001) and pharmacy ($102.40 versus $117.20; P <0.0001) costs. However, when central supply is put into consideration the total cost of coblation is slightly higher^27^ ($2,646 versus $2,591; P = 0.0011) with a difference of $55.94. However, because coblation is less likely to result in severe hemorrhage, it was found that using coblation tonsillectomy could save the National Health Service in the United Kingdom an incremental cost of £2000 for every avoided hemorrhage as opposed to using cold dissection with bipolar diathermy for haemostasis^28^. In a retrospective audit of 1,336 patients who had undergone coblation, it was reported that this technique had an increased requirement for operative intervention to manage secondary haemorrhage^29^.

This study supports the use of coblation technique over bipolar diathermy for pediatric patients undergoing tonsillectomy. To further support the current conclusion, the authors suggest using coblation tonsillectomy under rigorous and well-designed RCTs to appropriately record data for primary and secondary outcomes.

A systematic review and meta-analysis were performed on available studies in order to provide an evidence-based conclusion. The studies and their results were described and summarized, and the risk of bias in each study was appraised. The study designs and populations were standardized, with inclusion and exclusion criteria pre-defined. The interventions were homogenous across the included studies, with patients undergoing either coblation tonsillectomy or bipolar diathermy. Although some variation was observed in the control group such as using bipolar scissors as the primary cutting tool, bipolar diathermy was used as the primary technique for hemostasis. Despite these strengths, a number of inherit limitations should be considered. The current meta-analysis enrolled only seven studies with a total sample size of 1328 patients. There was a relatively large range in their sample sizes (1008 and 38 participants for Belloso et al. and Temple et al., respectively). Further clinical trials should therefore be completed to support the current study findings of the superior effect of coblation technique for pediatric patients undergoing tonsillectomy.

## Conclusions

Although the evidence is limited, with only seven studies comparing coblation and bipolar techniques, the results of this meta-analysis suggest that coblation improves the outcomes in paediatric patients undergoing tonsillectomy as it decreases delayed hemorrhage and post-operative pain and does not worsen intraoperative bleeding and reactionary hemorrhage. Further clinical studies are required to support the efficiency of coblation tonsillectomy technique.

## Data Availability

The datasets generated and analysed during the current study are available from the corresponding author on reasonable request.

## Declarations

### Ethics Approval and Consent to Participate

Not Applicable.

### Consent for Publication

Not Applicable.

### Competing Interests

The author(s) declared that they have no competing interests.

### Funding

The author(s) received no financial support for the research, authorship, and/or publication of this article.

### Author Contributions

Study concept and design: Mohammad Karam and Ahmad Abul

Acquisition of Data: Abdulredha Almuhanna, Abdulrahman AlNaseem, Abdulmalik Alsaif Analysis of Data: Mohammad Karam, Ahmad Abul, and Ali Aljadi

Interpretation of Data: Mohammad Karam, Ahmad Abul, Abdulwahab Althuwaini, Ali Aljadi, and Talal Alanezi

All authors contributed in the drafting of the manuscript.

Study Supervision: Athari Alwael

All authors have read and approved the final draft manuscript.

## Acknowledgements

Not Applicable.

## Notes

### Competing Interest Statement

The authors have declared no competing interest.

### Funding Statement

The authors received no financial support for the research, authorship, and/or publication of this article.

